# Leader and Follower Roles in Adapted Tango Reveal Complementary Neural Routes to Motor Improvement in Parkinson’s Disease

**DOI:** 10.64898/2026.09.14.26362964

**Authors:** Constantina Theofanopoulou, Neha Bajaj, Alberto Muñoz Sánchez, Bruce Crosson, Steven L Wolf, Venkatagiri Krishnamurthy, Keith M. McGregor, Madeleine E. Hackney

**Author notes:** Corresponding Authors: Constantina Theofanopoulou, 1200 York Ave, 10065, New York, NY, Madeleine E. Hackney, 57 Executive Park S, Suite 219, Atlanta, GA, 30329.

## Abstract

Parkinson’s disease (PD) disrupts internally generated (IG) movement through impaired striato-thalamo-cortical circuitry, while externally guided (EG) movement may recruit relatively preserved cerebello-thalamo-cortical pathways. We tested whether IG and EG movement strategies differentially shape clinical outcomes and resting-state functional connectivity by using partnered adapted tango as a naturalistic IG/EG movement manipulation. People with mild-to-moderate PD were randomized to 12 weeks of adapted tango as Leaders, adapted tango as Followers, or a non-dance health education Control condition. During therapeutic adaptango classes, Leaders self-initiated movement direction, timing, and amplitude, whereas Followers used auditory, tactile, and proprioceptive cues from the partner to guide their movement. We identified significant clinical improvement in both dance groups, with Leaders and Followers showing reduced overall disease severity and motor impairment, while Controls did not show a corresponding significant pattern. When Leaders and Followers were pooled, the Dance group also showed overall clinical and motor improvement, in addition to enhanced lower-limb motor outcomes. Connectivity findings suggested partly distinct neural routes to benefit. Leaders showed relative network stability and shifted directionally toward lower connectivity values resembling those of healthy older adults on cerebellar-sensorimotor connections that are typically strengthened in PD. Followers showed increased connectivity across sensorimotor and caudate-motor circuits, consistent with cue-supported action updating. Controls showed overlapping connectivity increases in some motor-network regions of interest, but without comparable clinical gains, suggesting that increased connectivity alone may not be adaptive. These findings indicate that adapted tango can improve motor outcomes in PD through complementary IG and EG movement-weighted mechanisms, with Leader training potentially supporting neural normalization and Follower training engaging distributed cue-to-action networks.

## Introduction

Parkinson’s disease (PD) is the second most prevalent age-associated progressive neurodegenerative disorder [1, 2]. Clinically, PD is defined by cardinal motor symptoms including rigidity, tremor, bradykinesia, and postural instability, which together impair movement initiation, execution, balance, and daily motor function [3–5]. These symptoms affect multiple motor domains: reduced movement amplitude and slowed initiation can disrupt gait, turning, postural transitions, reaching, hand dexterity, facial expression, and speech-related movements [4, 6, 7]. Thus, PD is not only a disorder of limb movement or walking, but constitutes a broad disruption of coordinated motor control across effectors, motivating rehabilitation approaches that target movement coordination across the whole body and its function rather than isolated symptoms alone.

One useful framework for understanding, and ultimately for rehabilitating, parkinsonian symptoms is the well-documented distinction between brain pathways supporting internally generated (IG) versus externally generated (EG) movements. IG movements, such as self-paced walking or spontaneous humming, are initiated without direct external cues and rely heavily on cortico-striatal circuits [8]. EG movements are triggered by external sensory stimuli, for example, stepping to a metronome or swaying to music [9–11], and are thought to engage relatively preserved cortico-cerebellar pathways in early-to mid-stage PD [8, 12–14]. Behavioral evidence supports this dissociation: people with PD (PwPD) often show greater deficits in IG than in EG movements, for instance, walking more smoothly when paced by an auditory cue than at a self-selected pace [15]. Neuroimaging work suggests a neural basis for this asymmetry: because dysfunction of the striato-thalamo-cortical (STC) circuit is central to PD pathophysiology, PwPD have particular difficulty with IG tasks, while EG movements, which likely rely on relatively spared neural regions, show activation patterns that differ little from those of controls, at least in earlier disease stages [16]. Under IG conditions, by contrast, the cerebello-thalamo-cortical (CTC) pathway appears to be recruited more heavily to accomplish motor tasks in people with PD than in neurotypical individuals, a pattern consistent with compensatory recruitment of EG-associated circuitry, which is comparatively robust to the dopaminergic deterioration that undermines STC pathways [8].

We recently tested this IG/EG framework directly using resting-state functional connectivity (rsFNC), comparing people with mild-to-moderate PD (while they were OFF-medication) — the same trial cohort analyzed here, at their pre-intervention baseline — to healthy older adults (HOA) (Theofanopoulou et al., 2026a) [17], using as regions of interest (ROIs) brain regions repeatedly associated with IG/EG movements and mapped onto the reproducible NeuroMark independent component (IC) template [18]. Primary motor cortext (M1) subregions corresponding to leg, hand, and larynx representations, and postcentral gyrus/primary somatosensory cortex (PoCG/S1), each showed increased connectivity with cerebellar territories — particularly Crus II and Lobules VIIIa/VIIIb — in people with PD relative to HOA, alongside increased postcentral gyrus-insula connectivity. The caudate nucleus, a core STC hub, showed the converse profile: increased connectivity with superior temporal gyrus but decreased connectivity with superior medial frontal gyrus and cerebellar Crus II, with reduced caudate–superior-medial-frontal-gyrus connectivity associated with greater disease severity. This pattern — cortico-cerebellar (EG-associated) hyperconnectivity alongside reduced cortico-striatal (IG-associated) coupling — provided direct rsFNC evidence for the compensatory-recruitment account above, and motivated our use of the same connectivity mapping here to test whether these circuits are modifiable by targeted movement training in people with PD.

Building on this circuit-level distinction, rehabilitation approaches for PD differ in which of these two systems they primarily train. Mobility training, partnered dance such as tango, non-partnered dance such as Dance for PD, tandem biking, and tai chi have each shown efficacy for improving motor function in PD [19–23], but vary in how heavily they use IG versus EG movement strategies, or combine the two. IG-weighted strategies emphasize increased focus on movement planning or preparation for self-initiated movement, helping PwPD approach average speeds and amplitudes of their movement by focusing on critical movement qualities such as length of speed of movements [24]. EG-weighted strategies encourage motor responses from external sensory cues, including visual, auditory, and tactile cues, which can improve clinically meaningful variables such as stride length and timing accuracy [25–28], cues may be especially relevant for partnered dance because they are processed rapidly and with relatively low attentional demand compared with visual or auditory cues [26, 27, 29, 30]. In practice, however, most rehabilitative movement therapies engage both striatal and cerebellar systems to some degree [22]. Therefore, isolating whether a given behavioral or neural benefit derives primarily from IG- or EG-weighted training remains a central methodological challenge.

To address this gap, we designed an experiment built around a single shared activity, partnered, adapted Argentine tango [31–34], that intrinsically involves two distinct roles with defined goals and rules: the leader and the follower. Adapted tango uses the embrace between two dance partners as a tactile conduit for passing movement information between them. Both roles draw on IG and EG systems: to dance together, dancers must entrain to shared auditory stimuli (the music) and to each other, and granted, the leader responds to environmental and partner cues. Critically, however, the two roles are not weighted equally between the IG and EG systems. The leader self-initiates the direction, timing, and amplitude of movement and therefore engages a comparatively IG-weighted strategy, likely drawing correspondingly more heavily on striatal circuits. The follower’s movement, by contrast, is continuously cued by proprioceptive input from numerous sensory channels — visual, auditory, and tactile cues transmitted through the partner embrace — placing relatively greater weight on an EG-weighted strategy that may engage cerebellar circuits more than striatal circuits [16]. Neither role activates one circuit exclusively, but comparing leader and follower roles within the same shared activity creates a within-paradigm gradient for isolating the relative contributions of training self-initiated versus externally cued movement strategies while holding constant potential confounders such as number of sessions, social environment, music, dance steps, and physical partnering.

In this study, we used this leader/follower tango paradigm to evaluate the relative behavioral and neural effects of IG-versus EG-weighted movement training in people with idiopathic, mild-to-moderate PD. Participants were randomized to a leader role, a follower role, or a non-dance interactive-learning control group and completed twenty 90-minute sessions over 12 weeks. We assessed clinical motor outcomes using the Movement Disorder Society-Sponsored Revision of the Unified Parkinson’s Disease Rating Scale (MDS-UPDRS) [35], including total score, Part III motor examination score, and effector-specific hand, leg, and larynx subscores, and measured rsFNC before and after the intervention. Primary analyses compared Leaders, Followers, and Controls in a 3-group framework. Because both dance roles were embedded within the same adapted tango intervention, we also ran a secondary 2-group analysis pooling Leaders and Followers into a single Dance group and comparing them against Controls.

For the connectivity analysis, we leveraged the same anatomically precise mapping strategy developed in our companion cross-sectional study where we compared rsFNC in PwPD and HOA [17]. Rather than relying on broad network labels, we mapped regions previously implicated in IG and EG movement pathways onto reproducible NeuroMark ICs [18] — discrete cortical and subcortical regions such as cerebellar Lobule VIIIb, dorsomedial precentral gyrus/M1, and lateral caudate. Because the PD sample in that cross-sectional study was the same cohort analyzed here at baseline, this shared IC framework allowed us to directly interpret the present trial’s connectivity findings against the disease-associated connectivity signature identified in the companion sample. This allowed us to ask not only whether leader and follower training changed connectivity, but also whether any changes moved toward or away from the pattern that distinguished this same group of people with PD from a group experiencing healthy aging.

We hypothesized that leading and following would produce partly distinct behavioral and neural profiles within a broader IG/EG motor-control network, with each role potentially weighting STC versus CTC loops differently, while treating these loops not as categorically or exclusively engaged. Behaviorally, if mobility gains are related primarily to training self-initiation, planning, and movement selection, then Leaders would demonstrate greater improvements than Followers or Controls, accompanied by a connectivity signature preferentially involving STC-related circuitry, or a combination of STC-related and additional motor-control loops. If mobility gains are related primarily to exploiting external cues during training, then Followers would demonstrate greater improvements than Leaders or Controls, accompanied by connectivity changes preferentially involving CTC-related circuitry, or a combination of CTC-related and additional sensorimotor/cue-integration loops. If both dance roles improve relative to Controls, this finding would suggest that adapted tango supports motor function through a combination of self-initiated planning, rhythmic entrainment, tactile cueing, and whole-body sensorimotor coordination, with corresponding neural effects potentially distributed across both STC and CTC loops, as well as potential additional circuits. Therefore, in addition to role-specific Leader-versus-Follower hypotheses, we hypothesized that the pooled Dance group would show greater clinical improvement than Controls.

## Methods

### Participants and Recruitment Criteria

Fifty-seven people with PD (57) were recruited through various channels, including the Atlanta Veterans Affairs (VA) Center for Visual and Neurocognitive Rehabilitation (CVNR) registry, the VA Informatics and Computing Infrastructure (VINCI) database, the Michael J. Fox Foundation website, the Movement Disorders Unit at Emory University, newsletters and support groups from Parkinson’s organizations, educational events, and word of mouth. Participants were selected based on the following inclusion criteria: all people with PD had received a clinical diagnosis of PD by a movement disorders specialist, in accordance with the United Kingdom Parkinson’s Disease Society Brain Bank diagnostic criteria [36]. Participants were required to be at least 40 years old and capable of walking 3 meters or more, with or without assistance. All participants met the criteria for undergoing an fMRI scan, including normal hearing (>40dB pure-tone threshold). PwPD in Hoehn and Yahr stages I–III were included [37]. Participants were excluded if they scored below 18 on the Montreal Cognitive Assessment (MoCA) [38, 39]. Other exclusion criteria included peripheral neuropathy, untreated major depression, a history of stroke, or traumatic brain injury. Depression was assessed using the Beck Depression Inventory-II (BDI-II) [40], and a score of ≥30, indicating severe depression, served as the exclusion threshold. Potential participants were required to have a unilateral onset of symptoms and demonstrate clear symptomatic improvement with antiparkinsonian medications such as levodopa. Participants with a tremor score greater than 1 on the Movement Disorders Society Unified Parkinson’s Disease Rating Scale (MDS-UPDRS) Part III in a lower limb or with moderate to severe head tremor, were excluded. Finally, all participants were tested in the OFF-medication state, defined as being more than 12 hours since their last dose of antiparkinsonian medication.

Participants were randomly assigned to one of three groups using a computer-generated randomization sequence (randomizer.org). Participants attended twenty, 90-minute adapted tango classes as a Leader or a Follower, or twenty 90-minute health education classes, within a 12-week period. Trained raters administered measures according to standard procedures. Table 1 shows the demographic and clinical characteristics of participants who completed all 20 lessons and both pre- and post-intervention assessments (Leaders, n=19; Followers, n=21; Controls, n=17). Regarding statistical analyses, for each continuous or ordinal variable, group means (± SD) were compared across the three groups using the Kruskal-Wallis test, consistent with the non-parametric framework used throughout the behavioral analyses in this study; sex distribution was compared using a chi-square test. Ethnicity is reported descriptively (n, %).

**Table 1.** Baseline demographic and clinical characteristics, by group. Values are mean ± SD for continuous/ordinal variables and n (%) for categorical variables. Group comparisons used the Kruskal-Wallis test for continuous/ordinal variables and a chi-square test for sex distribution; ethnicity is reported descriptively only. Sample sizes reflect participants who completed all 20 intervention sessions and both pre- and post-intervention assessments. **Abbreviations:** PD, Parkinson’s disease; SD, standard deviation; MoCA, Montreal Cognitive Assessment (score range 0-30, higher scores indicating better global cognitive function); Confidence in Physical Function (CPF) score (higher scores indicate greater confidence in physical function); Hoehn & Yahr stage, clinical staging of PD motor severity (range I-V, higher stage indicating greater severity); Recent falls, self-reported number of falls in the preceding 6 months.

| Variable | Leaders<br>(n=19) | Followers<br>(n=21) | Controls<br>(n=17) | p |
| --- | --- | --- | --- | --- |
| Sex, male, n (%) | 13 (68%) | 12 (57%) | 12 (71%) | .638 |
| Age, years | 69.5 ± 5.7 | 67.6 ± 8.9 | 66.6 ± 10.0 | .895 |
| Ethnicity, n (%) |  |  |  |  |
| White/Caucasian | 15 (79%) | 15 (71%) | 11 (65%) |  |
| Black/African-American | 3 (16%) | 4 (19%) | 4 (24%) |  |
| Multiracial | 1 (5%) | 1 (5%) | 1 (6%) |  |
| Other | 0 (0%) | 1 (5%) | 1 (6%) |  |
| Education, years | 16.5 ± 2.8 | 17.1 ± 2.3 | 16.4 ± 1.8 | .477 |
| Years since PD diagnosis | 6.2 ± 5.1 | 6.1 ± 5.0 | 6.4 ± 4.0 | .752 |
| Hoehn & Yahr stage | 2.1 ± 0.6 | 2.2 ± 0.7 | 2.2 ± 0.6 | .759 |
| MoCA total score (/30) | 25.6 ± 3.3 | 25.6 ± 3.4 | 26.6 ± 2.8 | .576 |
| Number of comorbidities | 3.6 ± 1.5 | 2.9 ± 1.8 | 3.9 ± 1.9 | .143 |
| Number of prescription medications | 6.3 ± 3.8 | 4.5 ± 2.7 | 6.1 ± 4.2 | .209 |
| Recent falls, number | 2.5 ± 3.7 | 10.4 ± 39.5 | 3.6 ± 8.1 | .624 |
| Composite Physical Function score (/24) | 20.5 ± 3.8 | 18.8 ± 6.0 | 18.4 ± 4.8 | .291 |

### Intervention: Adapted Tango (Leader and Follower Roles) and Non-Dance Educational Control

Participants were assigned to one of three groups: two groups danced adapted tango in either the leader or follower role, and the third (a non-dance control) attended health education sessions. Participants in the leader and follower groups attended twenty 90-minute adapted tango classes over a 12-week period. Participants danced exclusively in their assigned role for the full duration of the intervention. Adapted tango is a modified form of Argentine tango designed to target the movement impairments of individuals with PD through changes to the tango frame and steps [31, 34]. Composed of simple steps, tango involves frequent movement initiation and cessation, multi-directional perturbations, and varied rhythms. Participants were instructed to focus on trunk control and stepping strategies, coordination, kinesthesia, their partner, their trajectory, and the aesthetics of the steps. Class content followed a 20-class syllabus detailed in an adapted tango manual, which describes aging-specific motor impairments, fall risk and prevention, partnering enhancement, and rhythmic entrainment; this syllabus has previously been shown to be effective for teaching adapted tango to people with PD [34].

Class sizes were limited to 10 pairs of a participant with PD and a healthy dance partner to maximize safety. PD participants danced only with healthy partners, who rotated every 15-20 minutes, a widely used practice for enhancing learning in partnered dance instruction. Undergraduate and graduate students served as partners across all sessions. Within each class, participants danced only their assigned role, leader (IG) or follower (EG), for the entire 12-week intervention. Leaders determined the steps and the direction, timing, and amplitude of each successive movement. Followers attended to tactile pressure cues transmitted through their partner’s hands, palms, and forearms at their own elbows and forearms, using these cues to determine the direction, timing, and amplitude of each step [41]. Leaders and Followers heard identical music at the same time throughout each class, to control for any differential influence of auditory cueing between roles.

Participants assigned to the control group attended sessions of matched duration and frequency (twenty 90-minute sessions over 12 weeks) consisting of health education lectures. Each session comprised a 1-hour lecture followed by 30 minutes of interactive small-group and partnered discussion, structured to employ learning techniques that support memory retention; discussion was actively encouraged throughout the lecture itself. Lectures were delivered by medical students, faculty, and experts from the Emory School of Medicine and other local universities and organizations, covering current research on health topics relevant to an older adult population. Student volunteers moderated lectures, assisted with presentation of material, and led discussion sections. Participants in this group were instructed not to alter their habitual exercise routines for the duration of the study. Further detail on the control condition is available in Dillard et al. (2018) [42].

### Behavioral Outcome Measures and Statistical Analysis

We conducted behavioral assessments at two timepoints: pre-intervention, within 2 weeks of the first tango or health education session, and post-intervention, within 2 weeks of the final session. We ran behavioral analyses to evaluate clinical changes associated with the intervention. We focused our analyses on MDS-UPDRS outcomes, including MDS-UPDRS total score, MDS-UPDRS Part III motor score, and effector-specific subscores related to laryngeal, hand, and leg motor function. Only participants with usable rsfMRI data were included in the behavioral analyses to ensure direct correspondence between imaging and behavioral outcomes. In addition to the primary 3-group analyses, Leaders and Followers were pooled into a combined Dance group for secondary 2-group comparisons against Controls. Longitudinal analyses were performed using paired observations from participants with available scores at both timepoints for the relevant outcome measure.

In detail, primary behavioral outcomes included MDS-UPDRS total score (updrs_total) and UPDRS Part III motor score (updrs_3_sum). To examine whether intervention-related changes were preferentially associated with specific motor effectors, additional composite subscores were generated from selected MDS-UPDRS Part II and Part III items, as categorized in Theofanopoulou et al. (2026a) [17]. The larynx-related subscore included items associated with speech and voice function (updrs_2_1, updrs_2_2, updrs_2_3, updrs_2_4, updrs_3_1). The hand-related subscore included measures associated with upper limb motor control, dexterity, and tremor (updrs_2_7, updrs_3_3_b, updrs_3_3_c, updrs_3_4_a, updrs_3_4_b, updrs_3_5_a, updrs_3_5_b, updrs_3_6_a, updrs_3_6_b, updrs_3_15_a, updrs_3_15_b, updrs_3_16_a, updrs_3_16_b, updrs_3_17_a, updrs_3_17_b). The leg-related subscore included measures associated with lower limb movement, gait, posture, and lower extremity tremor (updrs_2_12, updrs_2_13, updrs_3_3_d, updrs_3_3_e, updrs_3_7_a, updrs_3_7_b, updrs_3_8_a, updrs_3_8_b, updrs_3_10, updrs_3_11, updrs_3_17_c, updrs_3_17_d). For each participant and session, subscore values were calculated as the sum of all available items within the corresponding category. Missing values were treated as missing at the item level rather than excluding participants from the full analysis, thereby retaining participants whenever sufficient paired data were available for the relevant outcome.

Behavioral analyses included four classes of comparisons: baseline between-group comparisons, post-intervention between-group comparisons, within-group pre-post comparisons, and between-group comparisons of change scores (difference-of-differences). Baseline comparisons were conducted to evaluate pre-intervention comparability across groups, whereas post-intervention comparisons assessed end-state divergence following the intervention period. Within-group analyses evaluated longitudinal behavioral change separately within each group, and difference-of-differences analyses assessed whether the magnitude of behavioral improvement differed between interventions by comparing participant-level change scores (Δ = post - pre). In addition to omnibus 3-group analyses (Leaders vs. Followers vs. Controls), direct pairwise comparisons between Leaders and Followers were conducted to evaluate potential differences between internally guided and externally guided dance training strategies.

Because several behavioral outcomes demonstrated non-normal distributions and sample sizes were modest, primary statistical inference relied on non-parametric testing. Kruskal-Wallis tests were used for omnibus 3-group comparisons, Mann-Whitney U tests were used for 2-group comparisons, and Wilcoxon signed-rank tests were used for within-group longitudinal analyses. Difference-of-differences analyses were performed using Mann-Whitney U tests applied to participant-level pre-post change scores. To reduce false positives arising from multiple testing, Benjamini-Hochberg false discovery rate (FDR) correction was applied jointly across all groups and outcomes within each analytic panel (e.g., across all 3-group within-group comparisons in a single family; across all 2-group Dance-vs-Control comparisons in a separate family). Following the thresholding framework used in the rsfMRI analyses, behavioral significance was evaluated primarily at a threshold of p < 0.01, and only results surviving both p < 0.01 and q < 0.05 are reported as significant.

### Resting-State Functional Magnetic Resonance Imaging Procedure

Participants underwent rsfMRI scanning at two timepoints: once before the intervention began within 2 weeks of the first tango or health education session) and once after completion of the 12-week intervention (within 2 weeks of the final session). Neuroimaging data were collected at the Center for Systems Imaging at Emory University using a 3T Siemens Trio scanner with a Siemens 12-channel head coil. Participants were instructed to lie still with their eyes closed for 9 minutes and 45 seconds. Foam padding was used to minimize head motion. Following the scan, participants were asked whether they had fallen asleep; if so, the scan was repeated. Resting-state blood oxygen level-dependent (BOLD) fMRI data were acquired using a standard echoplanar imaging (EPI) sequence with an iPAT acceleration factor of 2. The scan parameters were as follows: 55 contiguous 3 mm slices in the axial plane, interleaved slice acquisition, repetition time (TR) = 3000 ms, echo time (TE) = 24 ms, flip angle = 90°, bandwidth = 2632 Hz/pixel, field of view (FOV) = 230 mm, matrix = 76 x 76, and voxel size = 3.0 x 3.0 x 3.0 mm. The first three TRs were discarded to allow for scanner stabilization. An anatomical image was acquired using a high-resolution MPRAGE sequence with 176 contiguous sagittal slices, single-shot acquisition, TR = 2300 ms, TE = 2.89 ms, flip angle = 8°, FOV = 256 mm, matrix = 256 x 256, bandwidth = 140 Hz/pixel, and voxel size = 1.0 x 1.0 x 1.0 mm.

### Resting-State Functional Magnetic Resonance Imaging Data Preprocessing

The imaging data underwent initial quality checks by a single rater, followed by automated quality control using the Magnetic Resonance Imaging Quality Control (MRIQC) software. MRIQC evaluates data quality through various metrics, including signal-to-noise ratio, artifacts, spatial smoothness, and motion-related parameters, to identify potential issues that could affect subsequent analyses. This process flagged one dataset (sub131-ses01) as problematic due to quality control concerns, leading to its exclusion from the analysis. Framewise displacement (FD) was also computed for each resting-state dataset to quantify head motion. Preprocessing included slice-time correction and motion correction on the functional volumes using SPM12. Physiological noise from pulse and respiration was normalized to standard space. The first 8-10 scans were discarded to mitigate saturation effects. The Retrospective Image Correction (RETROICOR) algorithm was applied to remove physiological noise related to cardiac and respiratory fluctuations [43]. Cardiac and respiratory functions were monitored using photoplethysmography on the left index finger and a respiratory belt around the chest. BOLD signal fluctuations due to low-frequency cardiac and respiratory waveforms were detrended using established methods [44]. In addition, the GIFT pipeline implemented despiking via AFNI’s 3dDespike during the ICA stage to attenuate transient extreme signal outliers in voxel time courses. The corrected data were low-pass filtered (cutoff frequency: 0.1 Hz) to isolate low-frequency resting-state BOLD fluctuations (Cordes et al., 2001). EPI images were then spatially normalized to the Montreal Neurological Institute (MNI) 3mm isotropic template using nonlinear registration and smoothed with a 10 mm full-width half maximum (FWHM) Gaussian kernel. Signal intensities for each volume were z-transformed, excluding the first six volumes from the calculation of the mean and standard deviation to avoid pre-steady-state outliers.

### Regions of Interest selected within Internally Generated & Externally Generated Pathways

We selected the same 27 resting-state ROIs used in our companion cross-sectional study of PwPD and HOA [17], (Table 2) where these regions were identified through a systematic literature review of neuroimaging studies of IG and EG movement; full review methodology is reported in that study. To anchor these ROIs to reproducible functional units rather than approximate anatomical boundaries, each ROI was mapped to one or more independent components (ICs) using the NeuroMark pipeline [18], an automated independent component analysis (ICA) framework that identifies reproducible resting-state brain networks across large datasets and clinical populations. It does so by using previously validated component templates as priors, allowing subject-level component estimation to be anchored to stable large-sample network definitions. In some cases, a single ROI corresponded to one NeuroMark IC (e.g., SMA to IC84); in others, an ROI mapped to multiple replicable ICs within the same broader region, differing along anatomical axes such as the dorsoventral or mediolateral extent of a gyrus (e.g., PreCG/M1 mapped to three ICs — IC2, IC54, and IC66). The present study used this identical set of 27 IC-defined ROIs, enabling direct, IC-level comparison between the present trial’s connectivity findings and the disease-associated connectivity patterns identified in that companion sample.

**Table 2.**
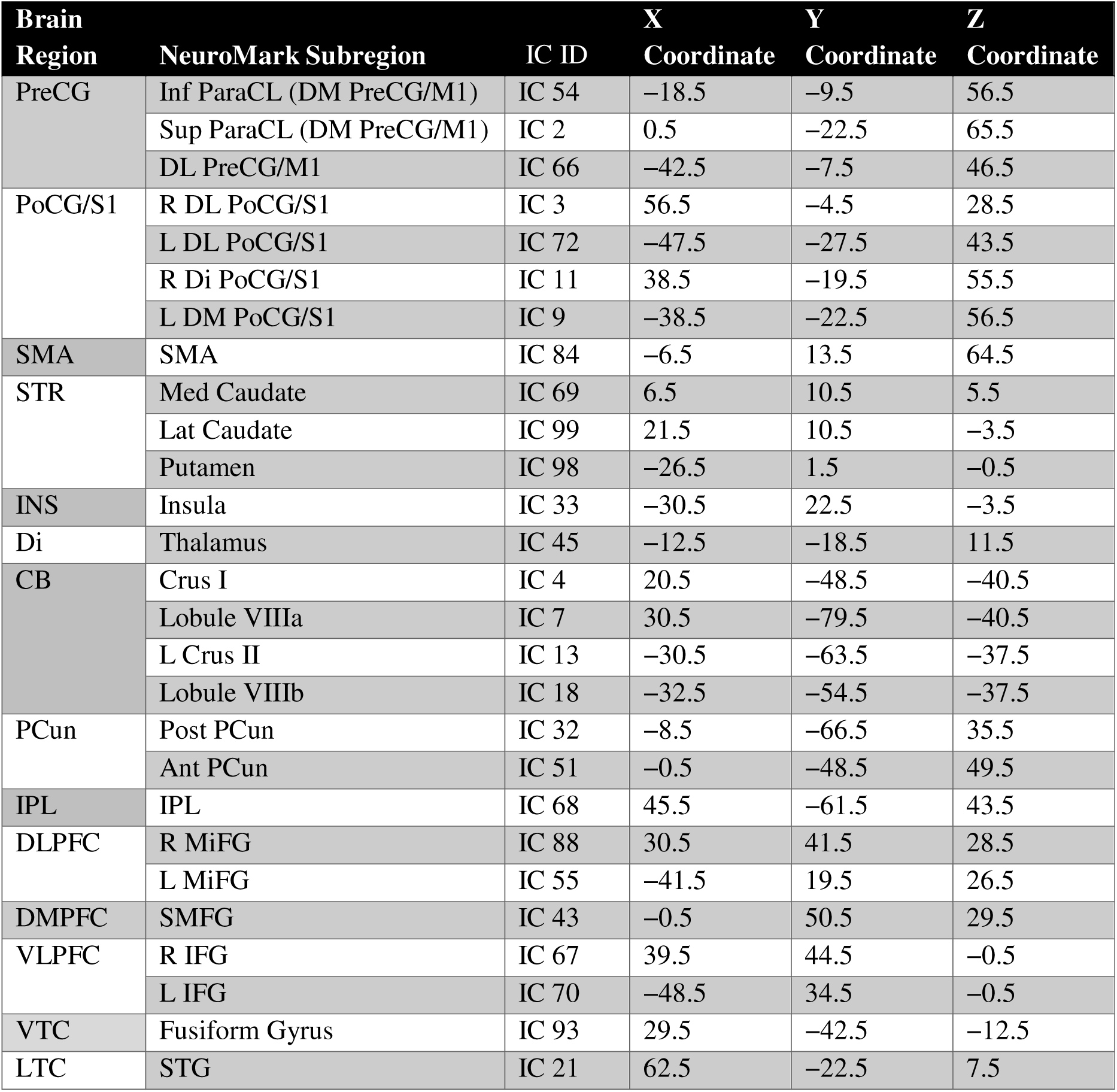
Peak MNI Coordinates of Selected NeuroMark Independent Components. This table shows the 27 NeuroMark ICs selected for analysis, which correspond to the ROIs repeatedly implicated in IG and EG neural pathways. Column 1 lists the broader brain region in which each NeuroMark IC is located. Column 2 specifies the subregion corresponding to each NeuroMark component. For regions without specific laterality indicated (i.e., R = Right or L = Left), the priors correspond to both hemispheres. Column 3 provides the index ID of each NeuroMark IC. Columns 4, 5, and 6 present the MNI X, Y, and Z peak coordinates for each NeuroMark component used in the analysis. Broader Brain Region **Abbreviations:** PreCG, Precentral Gyrus; SMA, Supplementary Motor Area; STR, Striatum; INS, Insula; Di, Diencephalon; CB, Cerebellum; PCun, Precuneus; IPL, Inferior Parietal Lobe; DLPFC, Dorsolateral Prefrontal Cortex; DMPFC, Dorsomedial Prefrontal Cortex; VLPFC, Ventrolateral Prefrontal Cortex; VTC, Ventral Temporal Cortex; LTC, Lateral Temporal Cortex. NeuroMark Subregion Abbreviations: Inf, Inferior; Sup, Superior; ParaCL, Paracentral Lobule; PreCG, Precentral Gyrus; M1, Primary Motor Cortex; DM, Dorsomedial; DL, Dorsolateral; Di, Dorsal-intermediate; PoCG, Postcentral Gyrus; SMA, Supplementary Motor Area; Med, Medial; Lat, Lateral; CB, Cerebellum; Post, Posterior; Ant, Anterior; IPL, Inferior Parietal Lobe; MiFG, Middle Frontal Gyrus; SMFG, Superior Medial Frontal Gyrus; IFG, Inferior Frontal Gyrus; STG, Superior Temporal Gyrus; Fusiform Gyrus (VTC).

Following preprocessing, the resulting time courses were analyzed using the GIFT software package (Group ICA of fMRI Toolbox) to perform spatially constrained independent component analysis (ICA) [45]. For each subject and session, the ICA procedure decomposed the fMRI time series into three primary outcome variables: a) component spatial maps: voxel-wise spatial representations of each independent component, illustrating the extent to which different brain regions contribute to a given network; b) component power spectra: frequency-domain information for each independent component, describing the spectral power distribution of the associated time series; and c) between-component connectivity (functional network connectivity, FNC): the temporal correlations between IC time courses used to quantify functional interactions between networks in all subsequent analyses.

### Statistical analysis of functional connectivity

All 27 ROIs, corresponding to NeuroMark ICs, were selected for analysis. FNC was quantified as the Pearson correlation between subject-specific independent component time courses obtained after ICA back-reconstruction. Correlation coefficients were Fisher z-transformed prior to group-level analysis. We conducted pairwise comparisons of FNC across all ICs were conducted, and the resulting spatial maps, power spectra, and submitted FNC matrices to group-level statistical analysis using the Multivariate Analysis of Covariance (MANCOVAN) toolbox, implemented in the latest version (as of August 2025) of the Group ICA of the fMRI Toolbox (GIFT). To evaluate group differences, independent t-tests were performed for all pairwise IC combinations for between-group comparisons (Leader vs. Follower, Leader vs. Control, Follower vs. Control). A significance threshold of p < 0.01 was applied to identify uncorrected effects. To mitigate false positives from multiple comparisons, false discovery rate (FDR) correction was performed using the MAFDR method [46], which estimates q-values as the minimum FDR threshold at which a test is considered significant. A threshold of q < 0.05 was used to determine FDR-corrected significance.

To comprehensively evaluate the effects of the intervention, we conducted four types of statistical comparisons, applied to both our primary 3-group framework (Leaders, Followers, Controls) and a secondary 2-group framework in which Leaders and Followers were pooled into a combined Dance group and compared against Controls. First, between-group comparisons at baseline assessed whether groups showed differences in connectivity before the intervention, establishing comparability across groups. Second, between-group comparisons post-intervention examined group differences in connectivity after the intervention, providing descriptive evidence of end-state divergence. Third, within-group post-pre comparisons tested for longitudinal connectivity change in each group separately, identifying networks altered by leader training, follower training, control participation, or dance participation overall. Finally, between-group difference-of-differences comparisons contrasted the magnitude of pre-to-post change across groups, offering the strongest test of whether the dance intervention produced connectivity changes beyond those attributable to time or non-specific factors.

### Connectivity Change Relative to the PwPD-vs-HOA Disease Signature

To assess whether trial connectivity findings could be interpreted in light of disease-related connectivity alterations, we mapped each significant resting-state functional network connectivity (rsFNC) pairwise result onto the previously published PwPD-vs-HOA cross-sectional analysis of the same 27-IC NeuroMark pipeline [17], in which the PwPD group was drawn from the same trial cohort analyzed here, at baseline. Because both studies used an identical 27×27 (351-pair) IC matrix, every IC pair we examined in this trial had a corresponding, previously computed PwPD-vs-HOA t-test result (Supplementary Table S2 of Theofanopoulou et al. 2026a) [17], regardless of whether that pair reached significance in the PwPD-vs-HOA comparison. For each IC pair, we determined (i) whether the identical pair appeared among the 19 IC pairs that survived the PwPD-vs-HOA significance threshold (p<0.01; 15 pairs of increased and 4 pairs of decreased connectivity in PwPD relative to HOA), and (ii) if so, the direction of the PwPD-vs-HOA effect. We classified IC pairs not meeting this threshold as showing no evidence of a disease-associated alteration at rest, independent of any change identified in this trial.

For the subset of IC pairs confirmed to correspond to a validated PwPD-vs-HOA disease signature, we examined each group’s (Leaders, Followers, Controls) connectivity trajectory relative to the independently acquired HOA reference sample (n=24, single timepoint). We treated the HOA sample as a fixed descriptive reference point against which each group’s own pre- and post-intervention values could be visually and directionally compared. For each comparison, we computed each group’s mean pre- and post-intervention FNC value and the within-subject change score (Δ = post - pre), with a one-sample t-test (Student’s t-distribution, n-1 degrees of freedom; SciPy) and 95% confidence interval on Δ to evaluate whether the change differed from zero within each group. To formally test whether the magnitude of change differed between groups, we fit a linear mixed-effects model for each of the four comparisons separately (FNC ∼ group × time, random intercept per subject, REML estimation; Control coded as the reference group and pre-intervention coded as the reference timepoint; implemented in Python’s statsmodels), yielding a group × time interaction coefficient, 95% CI, and p-value for the Leader-vs-Control and Follower-vs-Control contrasts at each edge. Across the 4 edges and 2 group contrasts (8 interaction tests total), we corrected resulting p-values for multiple comparisons using the Benjamini-Hochberg false discovery rate (FDR) procedure.

### Covariates

To evaluate whether rsfMRI or behavioral findings were associated with demographic or disease-related characteristics, we conducted exploratory covariate analyses including sex, age, ethnicity, and years since PD diagnosis, applied to our primary 3-group and secondary 2-group (Dance-pooled) frameworks.

For rsfMRI, we restricted covariate testing to the significant FNC pairs identified in the primary and secondary rsfMRI analyses above, rather than the full FNC matrix, to limit the multiple-comparison burden. For baseline and post-intervention FNC findings, we modeled the participant-level FNC value at the relevant session as a function of group and one covariate at a time. For within-group FNC changes and difference-of-differences FNC findings, we calculated ΔFNC values (post - pre) and modeled them as a function of group, where applicable, and one covariate at a time.

For behavioral outcomes, we calculated pre-post change scores (post - pre) for MDS-UPDRS total, MDS-UPDRS Part III, and the larynx-, hand-, and leg-related MDS-UPDRS subscores. For within-group analyses, we fit separate linear models within each group (Leaders, Followers, Controls, and the pooled Dance group) to test whether each change score was associated with one covariate at a time. For difference-of-differences analyses, we modeled change scores as a function of group and one covariate at a time, in both the 3-group and 2-group (Dance vs. Control) frameworks, to evaluate the covariate effect and the group effect after covariate adjustment.

We modeled sex and ethnicity as categorical predictors and age and years since diagnosis as continuous predictors, fitting models separately for each covariate to avoid overfitting given the sample size and sparse representation of some ethnicity categories. We evaluated statistical significance for exploratory covariate effects at uncorrected p<0.05, and applied Benjamini-Hochberg FDR correction across each analysis family, with q<0.05 considered significant after correction.

## Results

We conducted a randomized controlled trial in which PwPD were assigned to one of three 12-week interventions — leader-role adapted tango (Leaders), follower-role adapted tango (Followers), or non-dance health education (Controls) — with rsfMRI and MDS-UPDRS assessments collected before and after the intervention. We first confirmed that randomization produced comparable groups and that head motion did not differ across groups or timepoints, then evaluated intervention effects on rsFNC and clinical motor outcomes (MDS-UPDRS), tested whether either set of findings was explained by demographic or disease-duration covariates, and examined whether the trial’s connectivity findings could be interpreted in light of the disease-related connectivity signature identified in our companion cross-sectional study of PwPD versus HOA [47].

### Participant Characteristics

Randomization achieved balanced groups: Leaders, Followers, and Controls did not differ significantly on any demographic or baseline clinical characteristic tested, including age, sex, ethnicity, education, years since diagnosis, Hoehn & Yahr stage, MoCA score, comorbidity burden, medication count, or the Composite Physical Function score (**Table 1**).

### Head Motion

We assessed framewise displacement (FD) to confirm that group differences in connectivity could not be attributed to differential head motion while in the scanner. Mean FD did not differ between groups at baseline (Leaders: 0.203 ± 0.198 mm; Followers: 0.193 ± 0.114 mm; Controls: 0.186 ± 0.116 mm; Kruskal-Wallis p=.618) or post-intervention (Leaders: 0.134 ± 0.071 mm; Followers: 0.178 ± 0.220 mm; Controls: 0.165 ± 0.082 mm; p=.433). No group showed a significant within-group change in FD from pre- to post-intervention (Leaders p=.647; Followers p=.151; Controls p=.434), and the magnitude of FD change did not differ between groups (3-group difference-of-differences p=.800; Leader vs. Follower p=.643). We therefore interpret the connectivity findings below as unlikely to reflect differential head motion between groups.

### Resting-State Functional Connectivity

We evaluated rsFNC using four complementary comparisons, each addressing a distinct question. Baseline between-group comparisons tested whether groups differed in connectivity before the intervention, establishing pre-intervention comparability. Post-intervention between-group comparisons tested for connectivity differences after the intervention, providing descriptive evidence of end-state divergence between groups. Within-group pre-post comparisons tested for longitudinal connectivity change within each group separately, identifying networks altered by leader training, follower training, or control participation. Difference-of-differences comparisons directly contrasted the magnitude of pre-to-post change across groups, providing our strongest test of whether the dance intervention drove connectivity change beyond what would occur with time or non-specific factors alone. We applied this framework to our primary 3-group comparisons (Leaders, Followers, Controls) and, as a secondary analysis, to a 2-group comparison in which Leaders and Followers were pooled into a combined Dance group and compared against Controls. All reported effects survived at p<0.01 uncorrected; none survived MAFDR correction (q<0.05) (**Supplementary Tables S1-S4**).

#### Baseline between-group comparisons

We found few group differences at baseline. Followers showed lower connectivity than Leaders between dorsomedial PreCG/M1 and the anterior precuneus (p=.0039), and Controls showed lower connectivity than Leaders between left dorsolateral PoCG/S1 and the insula (p=.0067). These findings indicate that Followers and Controls began the intervention with reduced connectivity, relative to Leaders, in specific sensorimotor circuits (**Supplementary Table S1**).

In the pooled Dance-versus-Control comparison, we identified one additional baseline difference: the Dance group showed higher connectivity than Controls between medial caudate and Lobule VIIIb (p=.005), a difference not captured by the 3-group comparisons above (**Supplementary Table S1**).

#### Post-intervention between-group comparisons

We identified several differences post-intervention (**Figure 1**). Controls showed lower connectivity than Leaders between left dorsomedial PoCG/S1 and the insula (p=.0058), and higher connectivity than Leaders between left dorsomedial PoCG/S1 and cerebellar Lobule VIIIb (p=.0075) and between dorsomedial PreCG/M1 and cerebellar Lobule VIIIb (p=.0024). Controls also showed lower connectivity than Leaders between the insula and posterior precuneus (p=.0091) and between the insula and anterior precuneus (p=.0046), and higher connectivity than Leaders between right IFG and left IFG (p=.0074). Followers showed higher connectivity than Controls between left dorsolateral PoCG/S1 and MiFG (p=.0051). Together, these results indicate a post-intervention reorganization in which Controls showed both strengthened and weakened connectivity relative to Leaders across a distributed set of circuits, while Followers showed a more localized connectivity increase relative to Controls (**Supplementary Table S2**).

**Figure 1.**
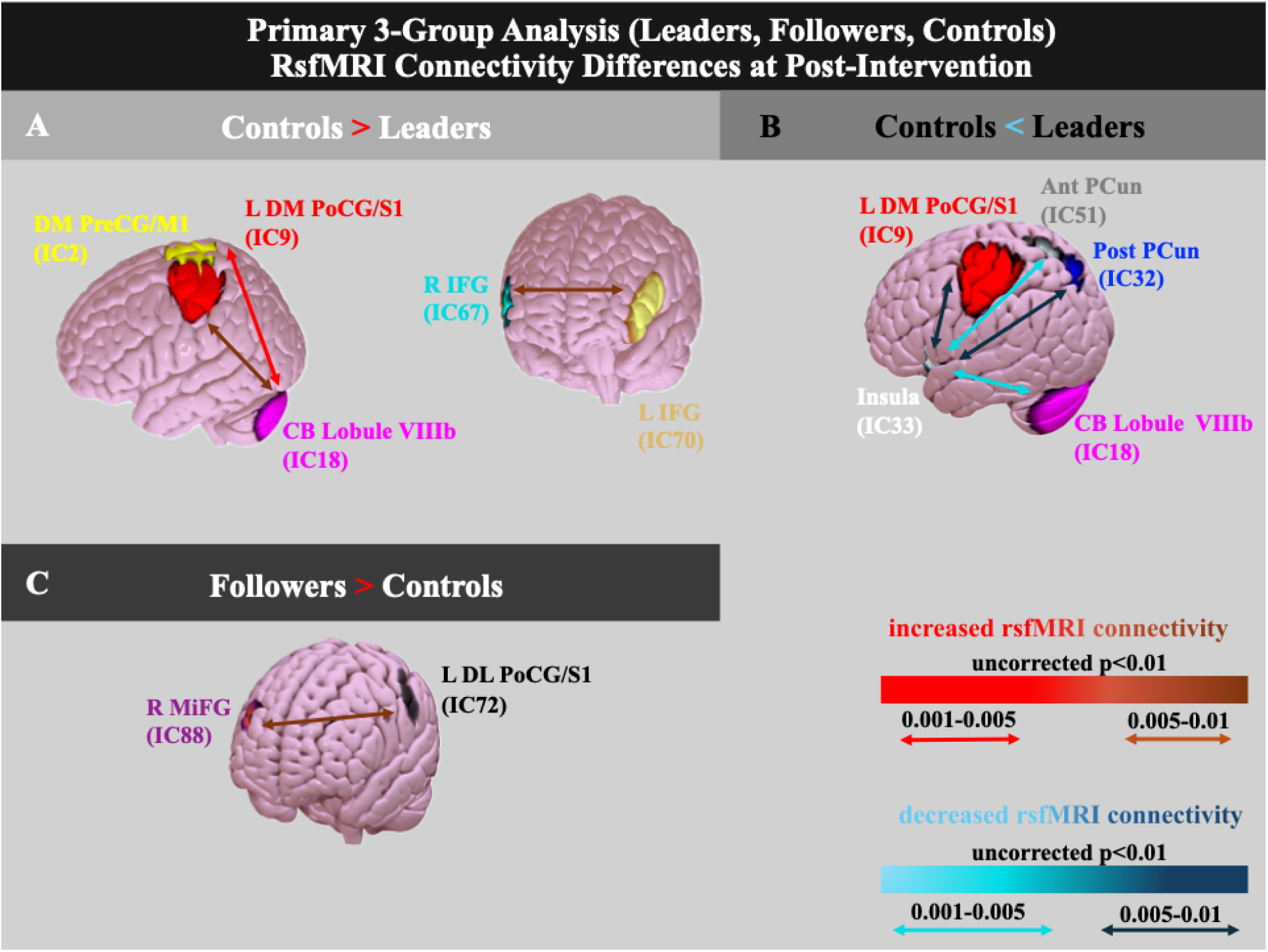
Primary 3-Group Analysis (Leaders, Followers, Controls). RsfMRI Connectivity Differences at Post-Intervention. **A. Controls > Leaders.** Increased functional connectivity in controls relative to leaders at post-intervention. B. Controls < Leaders. Decreased functional connectivity in controls relative to leaders at post-intervention. **C. Followers > Controls.** Increased functional connectivity in followers relative to controls at post-intervention. Color coding of connectivity arrows reflects significance levels from uncorrected t-tests (*p* < 0.01): red (p = 0.001-0.005), brown (p = 0.005-0.01). Increased connectivity is shown in warm colors; decreased connectivity is shown in cool colors. **Color coding and abbreviations:** IC9 (red): L DM PoCG/S1 (L: left; DM: dorsomedial; PoCG: postcentral gyrus; S1: primary somatosensory cortex); IC2 (yellow): DM PreCG/M1 (DM: dorsomedial; PreCG: precentral gyrus; M1: primary motor cortex); IC18 (fuchsia): CB Lobule VIIIb (CB: cerebellum); IC67 (cyan): R IFG (R: right; IFG: inferior frontal gyrus); IC70 (gold): L IFG (L: left; IFG: inferior frontal gyrus); IC33 (white): Insula; IC51 (gray): Ant Pcun (Ant: anterior; PCun: precuneus); IC32 (blue): Post PCun (Post: posterior; PCun: precuneus); IC88 (dark purple): MiFG (R: right; MiFG: middle frontal gyrus); IC72 (black): L DL PoCG/S1 (L: left; DL: dorsolateral; PoCG: postcentral gyrus; S1: primary somatosensory cortex).

In the pooled Dance-versus-Control comparison, we identified a broader set of post-intervention differences. The Dance group showed higher connectivity than Controls between the insula and Lobule VIIIb (p=.001), medial caudate and posterior precuneus (p=.002), medial Caudate and Lobule VIIIb (p=.004; which also differed at baseline, indicating this specific difference persisted rather than emerged during the intervention), SMA and left Crus II (p=.006), left IFG and posterior precuneus (p=.007), and lateral Caudate and posterior precuneus (p=.008). There was also lower connectivity than Controls between medial Caudate and insula (p=.008). These results indicate that the pooled Dance group diverged from Controls on a broader, largely caudate- and precuneus-centered set of regions not fully captured by the 3-group comparisons above (**Supplementary Table S2**).

#### Within-group pre-post comparisons

In our within-group pre-post comparisons, we found that Followers showed higher post-intervention connectivity between right dorsolateral PoCG/S1 and left dorsomedial PoCG/S1 (p=.0071) and between right dorsolateral PoCG/S1 and dorsolateral PreCG/M1 (p=.0090), indicating a strengthening of intra-sensorimotor and sensorimotor-motor cortex connectivity specifically following follower-role training. Leaders and Controls showed no within-group changes reaching this threshold (Supplementary Table S3).

When we pooled Leaders and Followers into a single Dance group, we identified five additional edges showing a nominal pre-to-post decrease, anatomically distinct from the increases identified in Followers alone: right dorsolateral PoCG/S1 with dorsolateral PreCG/M1 (p=.001) and with SMA (p=.001), left dorsomedial PoCG/S1 with dorsolateral PreCG/M1 (p=.001), right dorsal-intermediate PoCG/S1 with SMA (p=.002), and dorsolateral PreCG/M1 with SMA (p=.003) (**Supplementary Table S3**).

#### Difference-of-differences comparisons

Our difference-of-differences comparisons (**Figure 2**) revealed that Followers showed greater pre-to-post increases than Leaders in connectivity between the caudate and dorsomedial PreCG/M1 (p=.0048), right dorsolateral PoCG/S1 and MiFG (p=.0003), right dorsolateral PoCG/S1 and SMA (p=.0021), left dorsomedial PoCG/S1 and IPL (p=.0032), fusiform gyrus and cerebellar Lobule VIIIa (p=.0082), and SMFG and SMA (p=.0012). Controls showed greater increases than Leaders across an overlapping set of edges (caudate–dorsomedial PreCG/M1, p=.0073; right dorsolateral PoCG/S1-MiFG, p=.0005; right dorsolateral PoCG/S1-SMA, p=.0033; left dorsomedial PoCG/S1-IPL, p=.0050; SMFG-SMA, p=.0020), but without the fusiform-cerebellar effect unique to Followers. These results indicate that both Followers and Controls showed larger connectivity increases than Leaders across sensorimotor, premotor, and subcortical-cerebellar circuits, with Followers showing a marginally broader profile of change (**Supplementary Table S4**).

**Figure 2.**
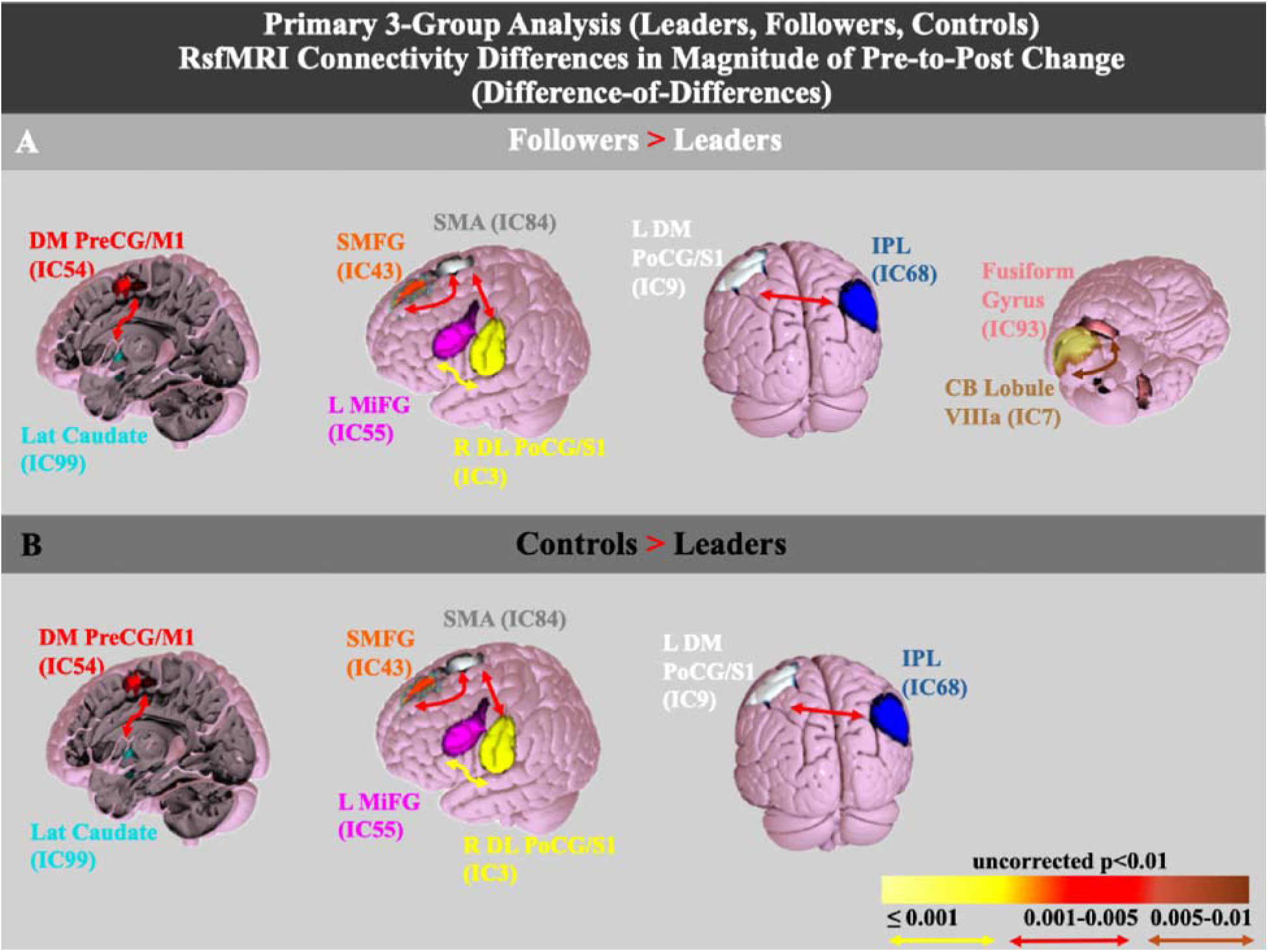
Primary 3-Group Analysis (Leaders, Followers, Controls). RsfMRI Connectivity Differences in Magnitude of Pre-to-Post Change (Difference-of-Differences). **A. Followers > Leaders.** Increased functional connectivity change (Δ = post - pre) in followers relative to leaders. **B. Controls > Followers.** Increased functional connectivity change (Δ = post - pre) in controls relative to followers. Color coding of connectivity arrows reflects significance levels from uncorrected t-tests (*p* < 0.01): yellow (p ≤ 0.001), red (p = 0.001-0.005), brown (p = 0.005-0.01). **Color coding and abbreviations:** IC54 (red): DM PreCG/M1 (DM: dorsomedial; PreCG: precentral gyrus; M1: primary motor cortex); IC99 (cyan): Lat Caudate (Lat: Lateral); IC55 (fuchsia): L MiFG (L: lateral; MiFG: middle frontal gyrus); IC3 (yellow): R DL PoCG/S1 (R: right; DL: dorsolateral; PoCG: postcentral gyrus; S1: primary somatosensory cortex); IC84 (gray): SMA (SMA: supplementary motor area); IC43 (orange): SMFG: superior medial frontal gyrus; IC9 (white): L DM PoCG/S1 (L: left; DM: dorsomedial; PoCG: postcentral gyrus; S1: primary somatosensory cortex); IC68 (blue): IPL (IPL: inferior parietal lobule); IC93 (pink): Fusiform Gyrus; IC7 (gold): CB Lobule VIIIa (CB: cerebellum).

**Figure 3.**
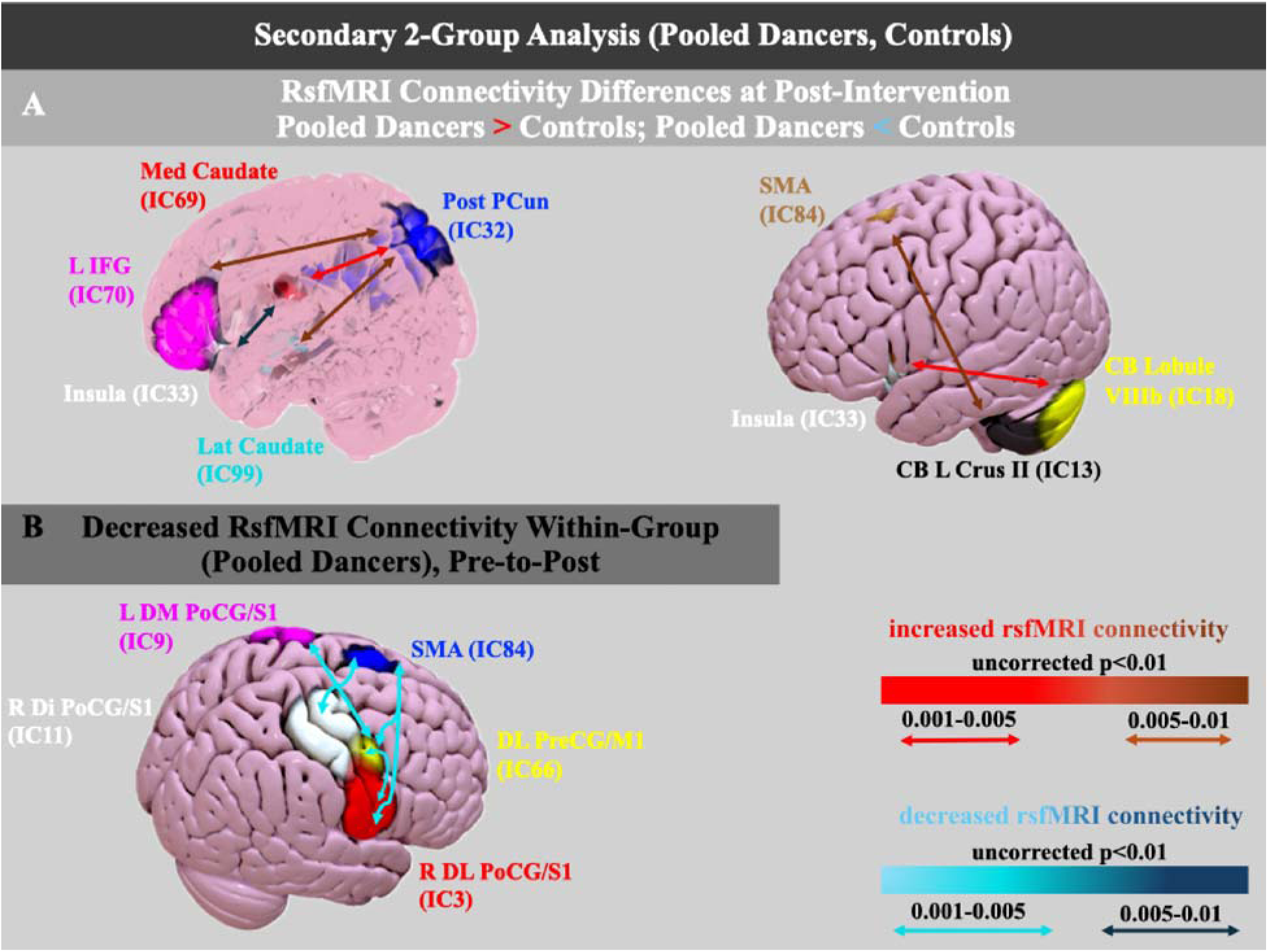
Secondary 2-Group Analysis (Pooled Dancers, Controls). **A. RsfMRI Connectivity Differences at Post-Intervention: Pooled Dancers > Controls; Pooled Dancers < Controls.** Increased functional connectivity in pooled dancers relative to controls at post-intervention, with one exception: the Medial Caudate-Insula edge shows decreased functional connectivity in pooled dancers relative to controls. **Color coding and abbreviations, panel (a):** IC69 (red): Med Caudate (Med: medial); IC32 (blue): Post PCun (Post: posterior; PCun: precuneus); IC70 (fuchsia): L IFG (L: left; IFG: inferior frontal gyrus); IC33 (white): Insula; IC99 (cyan): Lat Caudate (Lat: lateral); IC84 (gold): SMA (supplementary motor area); IC18 (yellow): CB Lobule VIIIb (CB: cerebellum); IC13 (black): CB L Crus II (CB: cerebellum; L: left). **B. Decreased RsfMRI Connectivity Within-Group (Pooled Dancers), Pre-to-Post.** Decreased functional connectivity within the pooled dance group from pre- to post-intervention. **Color coding and abbreviations, panel (b):** IC9 (fuchsia): L DM PoCG/S1 (L: left; DM: dorsomedial; PoCG: postcentral gyrus; S1: primary somatosensory cortex); IC84 (blue): SMA (supplementary motor area); IC11 (white): R Di PoCG/S1 (R: right; Di: dorsal-intermediate); IC66 (yellow): DL PreCG/M1 (DL: dorsolateral; PreCG: precentral gyrus; M1: primary motor cortex); IC3 (red): R DL PoCG/S1 (R: right; DL: dorsolateral).Color coding of connectivity arrows reflects significance levels from uncorrected t-tests (p < 0.01): red (p = 0.001-0.005), brown (p = 0.005-0.01) for increased connectivity; light blue (p = 0.001–0.005), dark blue (p = 0.005-0.01) for decreased connectivity.

**Figure 4.**
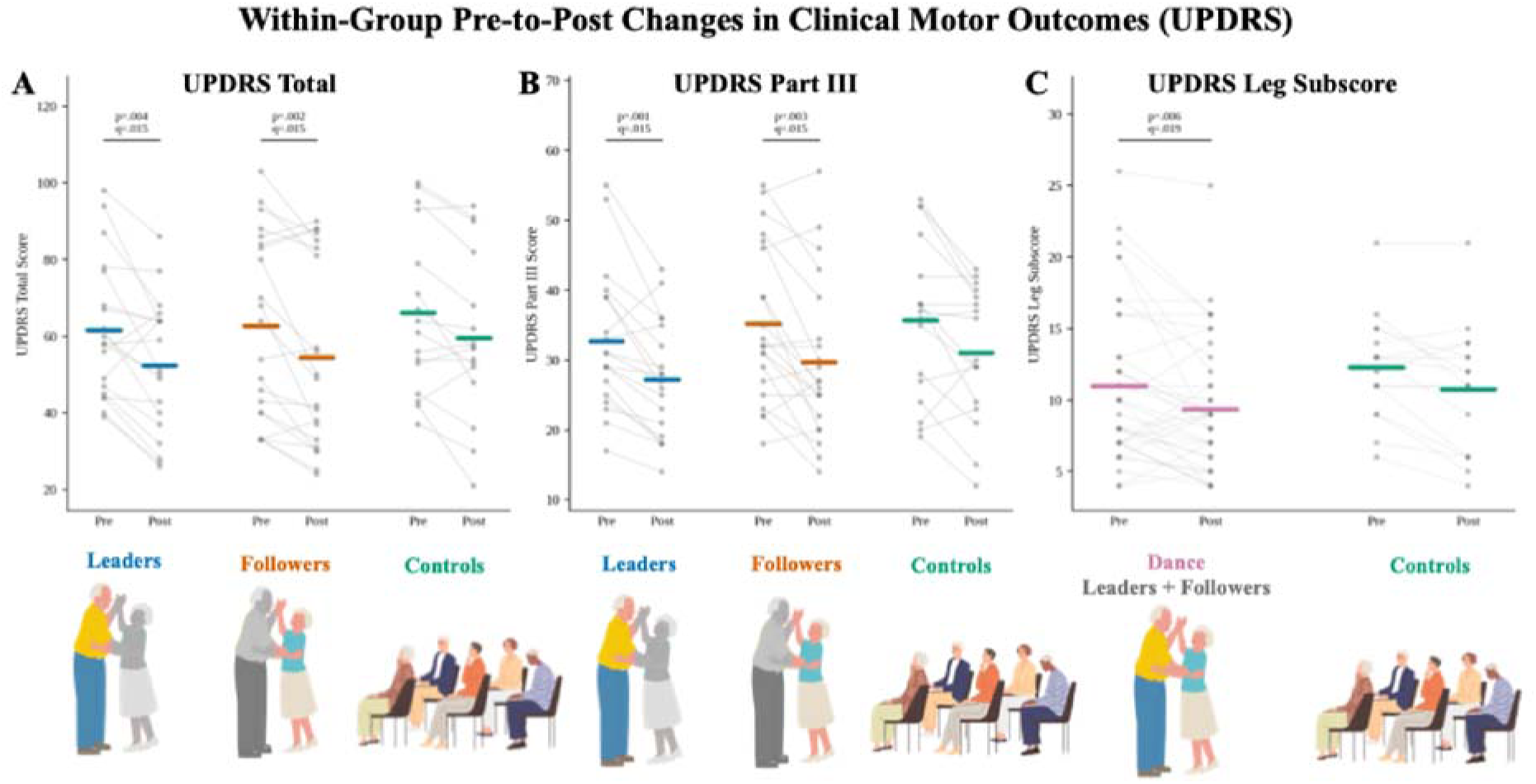
Within-Group Pre-to-Post Changes in Clinical Motor Outcomes (MDS-UPDRS). **A. MDS-UPDRS Total Score, Primary 3-Group Analysis (Leaders, Followers, Controls).** Individual participant trajectories (gray lines and points) and group means (colored horizontal bars) at pre- and post-intervention for UPDRS total score. **B. MDS-UPDRS Part III Score, Primary 3-Group Analysis (Leaders, Followers, Controls).** Individual participant trajectories and group means at pre- and post-intervention for UPDRS Part III (motor examination) score. **C. MDS-UPDRS Leg Subscore, Secondary 2-Group Analysis (Pooled Dancers, Controls).** Individual participant trajectories and group means at pre- and post-intervention for the leg-related UPDRS subscore, with Leaders and Followers pooled into a single Dance group. Each gray line connects one participant’s pre- and post-intervention score; darker gray points indicate overlapping participant values. Colored horizontal bars indicate the group mean at each timepoint. Brackets and accompanying p- and q-values denote within-group Wilcoxon signed-rank tests, Benjamini-Hochberg FDR-corrected within each analytic panel; only comparisons meeting the pre-specified dual threshold of p<0.01 and q<0.05 are annotated. Comparisons not meeting this threshold (Controls in panels A and B; Controls in panel C) are shown without brackets. **Color coding and abbreviations:** Blue: Leaders. Orange: Followers. Teal: Controls. Pink: Dance (Leaders and Followers pooled). Gray: individual participant data. UPDRS: Unified Parkinson’s Disease Rating Scale.

In the pooled Dance-versus-Control comparison, we found no pair of regions reaching this threshold, indicating that the pooled Dance and Control groups did not differ in the magnitude of pre-to-post connectivity change on any edge.

### Behavioral (UPDRS) Outcomes

We evaluated clinical motor outcomes using the same four-comparison framework applied to rsFNC: baseline group differences, post-intervention group differences, within-group pre-to-post change, and between-group differences in the magnitude of change. We applied this framework to MDS-UPDRS total score, MDS-UPDRS Part III, and effector-specific subscores indexing hand-, leg-, and larynx-related motor function. Primary analyses compared Leaders, Followers, and Controls, while secondary analyses pooled Leaders and Followers into a combined Dance group for comparison with Controls. We corrected for multiple comparisons within each analytic panel and report as significant only effects meeting the pre-specified dual threshold of p<0.01 and q<0.05. We additionally conducted an exploratory item-level analysis across individual UPDRS items to determine whether composite-level effects were driven by specific motor features.

#### Baseline between-group comparisons

We found no significant group differences on any behavioral outcome at baseline (all Kruskal-Wallis p>0.01; all q>0.05; **Supplementary Tables S5, S6**), confirming comparable pre-intervention clinical severity.

#### Post-intervention between-group comparisons

Post-intervention, groups again did not differ significantly after correction (all q>0.05; **Supplementary Table S7)**, including direct Leader-versus-Follower comparisons (all Mann-Whitney U p>0.01; all q>0.05), although Dance participants showed lower post-intervention MDS-UPDRS scores than Controls.

#### Within-group pre-post comparisons

Within-group comparisons revealed the clearest behavioral effects (**Supplementary Table S8**). Leaders showed significant reductions in UPDRS total (p=0.004, q=0.015) and UPDRS Part III (p=0.001, q=0.015). Followers showed significant improvements in UPDRS total (p=0.002, q=0.015) and UPDRS Part III (p=0.003, q=0.015). When we pooled Leaders and Followers into a single Dance group, both UPDRS total and UPDRS Part III remained significant (both p<0.001, both q<0.001); the Dance group additionally showed a significant reduction in the leg-related subscore (p=0.006, q=0.019). Controls did not meet this dual threshold on any global or effector-specific outcome.

#### Difference-of-differences comparisons

Between-group comparisons of change scores did not survive correction (all q>0.05; **Supplementary Table S9**), in either the omnibus 3-group analysis or the direct Leader-versus-Follower comparison. Dance participants showed numerically larger improvements than Controls, and Followers showed numerically greater improvement than Leaders on some hand-related measures, but these differences remained modest at the level of gross clinical UPDRS scores.

#### Exploratory item-level analyses

To determine whether these effects were driven by specific individual items, we repeated this framework across all 63 individual UPDRS items (**Supplementary Tables S10-S14)**; given the number of tests, we treated these findings as exploratory. We found no corrected group-specific effects in any item at baseline, post-intervention, or difference-of-differences item-level analyses, and no item in any of these three comparisons reached even the nominal significance p<0.01 threshold (Supplementary Tables S11, S12, S14).

Within-group, several items showed nominal (p<0.01, uncorrected) pre-post change, but only one survived FDR correction (**Supplementary Table S13**). In the 3-group framework, Leaders showed nominal improvement in speech (updrs_3_1; p=0.0039, q=0.147), arising from chair (updrs_3_9; p=0.0075, q=0.158), and gait (updrs_3_10; p=0.0047, q=0.147), and Controls showed nominal improvement in arising from chair (updrs_3_9; p=0.0067, q=0.358); none of these survived correction. In the 2-group framework, the pooled Dance group showed nominal improvement in fatigue (updrs_1_13; p=0.0027, q=0.074), speech (updrs_3_1; p=0.0067, q=0.074), facial expression (updrs_3_2; p=0.0043, q=0.074), arising from chair (updrs_3_9; p=0.0061, q=0.074), and body bradykinesia (updrs_3_14; p=0.0070, q=0.074), none of which survived correction, and gait (updrs_3_10; p=0.00016, q=0.010), which did survive FDR correction. Controls again showed a nominal improvement in arising from chair (updrs_3_9; p=0.0067, q=0.358) that did not survive correction; given that this item showed no supporting cluster of related nominal changes across either framework and occurred at a magnitude consistent with the expected false-positive rate across 63 comparisons, we interpret it as more likely attributable to chance or test-retest effects than to a genuine Control-specific change.

### Covariate Analyses

Although randomization achieved balanced groups on all baseline demographic and clinical characteristics tested (**Table 1**), we conducted exploratory covariate analyses to test whether sex, age, ethnicity, or years since diagnosis nonetheless predicted the magnitude of connectivity or behavioral change, independent of group membership, in our primary 3-group and secondary 2-group frameworks (**Supplementary Table S15)**.

For the connectivity findings, no covariate significantly predicted any of the significant FNC pairs identified in the primary 3-group analysis after correction, and the FNC group effects reported above remained significant after covariate adjustment. We found the same pattern for the significant FNC pairs identified in the secondary 2-group (Dance-vs-Control) analysis: no covariate survived FDR correction (all q>0.59), and the Dance-vs-Control group effect remained significant after covariate adjustment for every edge tested (all p<011).

For the behavioral findings, no covariate effect survived FDR correction in either the 3-group or 2-group frameworks (all q>0.05). We observed a nominal association between age and larynx-related change in Controls (0=0.167, p=0.002, q=0.102), such that older age was associated with less improvement (or greater worsening) in larynx-related scores, but this did not survive correction. We observed similar nominal age effects in larynx-related difference-of-differences models (3-group: 0=0.090, p=0.014, q=0.517; Dance vs. Control: 0=0.087, p=0.017, q=0.517), again not surviving correction, with the corresponding group comparisons remaining non-significant. Within the pooled Dance group, no covariate reached even nominal significance for any within-group behavioral change score (smallest: years since diagnosis on hand subscore, p=.180). These analyses indicate that the principal behavioral and connectivity findings, in both the 3-group and 2-group frameworks, are not explained by the demographic or disease-duration variables tested here.

### Connectivity Change Relative to the PwPD-vs-HOA Disease Signature

In our companion paper, rsFNC in PwPD was compared against HOA using the same 27 regions of interest and identical scanning and preprocessing framework applied here; the PwPD in that comparison are the same individuals who went on to enroll in the present trial, at their pre-intervention baseline. Because both studies used this identical set of regions, we were able to ask whether any of the present trial’s own significant connectivity findings corresponded to the specific connections that had already been shown to distinguish PD from healthy aging. To do this, we checked each significant connectivity result from the present trial against the full pairwise connectivity comparison from that companion study, and determined whether it matched one of the connections independently identified there as differing between PwPD and HOA (**Table 3**).

**Table 3.** Trial rsFNC findings validated against the independent PwPD-vs-HOA disease-connectome signature. This table lists the four IC-pair edges, among the trial’s significant rsFNC findings, that are also part of the disease-connectome signature independently validated in the companion PwPD-vs-HOA cross-sectional study (same NeuroMark 27-IC pipeline; full mapping in Supplementary Table S16). “Direction in PwPD vs. HOA” reflects that independent study, not the present trial. The Leader vs. Control and Follower vs. Control columns report group × time mixed-effects model coefficients and Benjamini-Hochberg FDR-corrected q-values from the present trial, Control coded as the reference group (full model output in Supplementary Table S17). **Abbreviations:** IC = independent component; PreCG = precentral gyrus; M1 = primary motor cortex; PoCG = postcentral gyrus; S1 = primary somatosensory cortex; CB = cerebellum; PwPD = people with Parkinson’s disease; HOA = healthy older adults; FDR = false discovery rate; 0 = model coefficient; q = FDR-corrected p-value.

| Edge | Trial contrast(s) | Direction in PwPD vs. HOA | Leader vs. Control ($\beta$, q) | Follower vs. Control ($\beta$, q) |
| --- | --- | --- | --- | --- |
| PreCG/M1 (IC2) - CB Lobule VIIIb (IC18) | Controls > Leaders, post (3-group) | Increased in PwPD | -0.162, q=.40 | 0.033, q=.84 |
| L DM PoCG/S1 (IC9) - CB Lobule VIIIb (IC18) | Controls > Leaders, post (3-group) | Increased in PwPD | -0.185, q=.35 | -0.094, q=.64 |
| L DM PoCG/S1 (IC9) - Insula (IC33) | Leaders > Controls, post (3-group) | Increased in PwPD | 0.251, q=.35 | -0.031, q=.84 |
| Insula (IC33) - CB Lobule VIIIb (IC18) | Leaders/Dance > Controls, post (3-group and 2-group) | Decreased in PwPD | 0.215, q=.35 | 0.196, q=.35 |

Of the present trial’s 23 significant connectivity findings across all 3-group comparisons, four IC pairs corresponded to pairs independently validated as disease-related in that prior analysis: PreCG/M1-CB Lobule VIIIb, left dorsomedial PoCG/S1-CB Lobule VIIIb, and left dorsomedial PoCG/S1-Insula, all elevated in PwPD relative to HOA, and Insula-CB Lobule VIIIb, reduced in PwPD relative to HOA. The remaining findings from the present trial corresponded to IC pairs that did not differentiate PwPD from HOA in the reference dataset and are therefore not attributable to a known disease-connectivity signature as identified in our companion study (**Supplementary Table S16**).

We applied this same mapping procedure to the secondary 2-group (Dance-vs-Control) findings reported above. Of the 13 significant Dance-vs-Control and within-Dance edges, only one corresponded to a validated disease signature: the Dance group’s higher post-intervention connectivity between the insula and cerebellar Lobule VIIIb (the same pair already identified in the 3-group framework above), indicating this signal persists when Leaders and Followers are pooled. The remaining 12 pairs, including the entire within-Dance pre-to-post pattern, corresponded to IC pairs that did not differentiate PwPD from HOA in the reference dataset (**Supplementary Table S16**).

Restricting to the four validated pairs from the 3-group framework, we asked whether each group’s connectivity moved toward or away from the independent HOA reference sample. For each edge, we compared each group’s mean pre- and post-intervention connectivity value to the fixed HOA reference value at that same edge, computed each group’s within-subject change (Δ = post - pre) with a one-sample t-test and 95% confidence interval, and additionally fit a linear mixed-effects model (connectivity ∼ group × time) to formally test whether the magnitude of change differed between groups. Leaders showed a directionally consistent, though not statistically significant, shift toward the HOA reference on three of four pairs (largest: PreCG/M1-CB Lobule VIIIb, Δ=-0.134, 95% CI [-0.288, 0.021], p=.085), while Controls showed the largest single-edge shift in the entire dataset away from the HOA reference (Insula-CB Lobule VIIIb, Δ=-0.181, 95% CI [-0.410, 0.048], p=.114). Followers also showed a non-significant shift toward the HOA reference on three of four pairs, most notably on left dorsomedial PoCG/S1-Insula (Δ=-0.162, 95% CI [-0.405, 0.080], p=.177), but moved away from the HOA reference on PreCG/M1-CB Lobule VIIIb (Δ=+0.061, 95% CI [-0.067, 0.190], p=.329), the same pair on which Leaders showed their strongest shift toward HOA. Formal group×time mixed-effects models confirmed this pattern numerically, but no contrast survived FDR correction across the 8 group comparisons tested (all FDR pA.35). This exploratory analysis of the four connectivity edges independently validated as PwPD-vs-HOA disease markers therefore revealed a directionally consistent, but not statistically significant, pattern of change across groups (**Supplementary Table S17**).

## Discussion

This study was designed to determine whether IG-weighted or EG-weighted movement training produces differential engagement of brain pathways and/or differences in clinical outcomes and motor performance in people with mild-to-moderate PD, when compared to a non-dance education control group. Leaders, who self-initiate the direction, timing, and amplitude of each movement, practiced a comparatively IG-weighted strategy. Followers, whose movements were continuously cued through music and the partner embrace, practiced a comparatively EG-weighted strategy. Because both roles were embedded within the same partnered dance intervention, and because an important clinical question is whether dance-based training benefits PwPD regardless of role, we additionally ran a secondary analysis pooling Leaders and Followers into a single Dance group and comparing them against Controls.

Three findings anchor the interpretation that follows. First, Leaders’ rsFNC was comparatively stable across the intervention: no IC pair showed a significant within-group change in Leaders, and no difference-of-differences result identified Leaders as showing more change than either Followers or Controls. Second, Followers showed a small number of significant within-group increases in sensorimotor connectivity and, together with Controls, showed significantly larger pre-to-post increases than Leaders across a substantially overlapping set of caudate-, sensorimotor-, premotor-, and parietal-related edges. Third, despite this apparent similarity between Followers and Controls in the difference-of-differences results, the two groups diverged clinically: Followers, like Leaders, showed statistically significant improvement on both UPDRS total and UPDRS Part III scores, whereas Controls did not show significant improvement. These findings suggest that connectivity stability and connectivity change can both be compatible with clinical improvement, but may reflect different mechanisms. Leaders improved clinically in the context of relative network stability and possible movement toward HOA-like values on disease-relevant cerebellar-sensorimotor pairs. Followers improved clinically in the context of increased sensorimotor and caudate-motor connectivity, consistent with cue-supported sensorimotor updating. Controls showed some connectivity increases similar to Followers, but without robust clinical associations, suggesting that increased connectivity alone should not be interpreted as inherently adaptive.

### Interpreting rsFNC change through the IG/EG lens

We interpret these patterns primarily through the IG/EG lens that motivated the trial design, while treating the broader normalization-versus-compensation framework from the PD connectivity literature as supporting context. Prior neuroimaging studies have shown that IG and EG movements recruit partially overlapping but differentially weighted motor circuits. IG movements rely heavily on cortico-striatal and medial motor systems, including caudate/putamen, SMA/pre-SMA, M1, and associated frontal motor-planning regions. EG movements draw strongly on sensorimotor and cue-integration systems, including M1, PoCG/S1, cerebellar lobules, IPL, premotor/frontal regions, and, depending on task demands, insula and precuneus [8, 12, 16, 48–53]. This literature provides the basis for asking whether Leader and Follower roles, while embedded in the same adapted tango intervention, differentially weight STC- and CTC-related pathways.

Under this lens, Leaders’ comparative connectivity stability is consistent with a preserved or stabilized IG-weighted operating point. Leaders, who in their dancing role may engage networks relevant to IG movement planning, did not show widespread rsFNC increases over time in IG-weighted or other networks. Instead, the clearest disease-signature context emerged from the additional exploratory analysis in which we compared the direction of connectivity change in the present trial with our companion study, where the same PwPD that were later split into the three intervention arms here were contrasted against HOA. Leaders showed a directionally consistent shift toward HOA reference values, specifically toward decreased PreCG/M1-cerebellar Lobule VIIIb and PoCG/S1-cerebellar Lobule VIIIb connectivity, whereas Controls showed greater post-intervention connectivity than Leaders on these disease-elevated cerebellar-sensorimotor pairs. This pattern is compatible with a cautious remediation account: Leader training may have supported clinical improvement without requiring broad compensatory up-regulation of motor-network connectivity, whereas the Control condition may have been associated with persistence or accentuation of a disease-elevated cerebellar-sensorimotor pattern. This interpretation should remain exploratory, but it provides a coherent link between the trial and the companion disease-signature analysis.

Followers showed a different neural profile. In the within-group analysis, they showed increased connectivity between right dorsolateral PoCG/S1 and left dorsomedial PoCG/S1, and between right dorsolateral PoCG/S1 and dorsolateral PreCG/M1. These findings point to strengthened intra-sensorimotor and sensorimotor-motor connectivity, rather than isolated cerebellar recruitment. In the difference-of-differences analysis, Followers showed greater pre-to-post increases than Leaders in caudate-dorsomedial PreCG/M1, right dorsolateral PoCG/S1-MiFG, right dorsolateral PoCG/S1-SMA, left dorsomedial PoCG/S1-IPL, fusiform gyrus-cerebellar Lobule VIIIa, and SMFG-SMA. This pattern is consistent with the literature showing that externally cued or rhythmically structured movement recruits not only cerebellum but also M1, S1, SMA, IPL, striatum, and frontal regions [12, 48, 49, 51, 53–56]. Therefore, the Follower profile is better understood as a multisensory sensorimotor-updating signature, with one cerebellar contribution, rather than as a simple CTC-only effect.

This pattern refines the initial EG/CTC hypothesis. The Follower role is EG-weighted because Followers receive auditory input from music and continuous tactile/proprioceptive input through the partner embrace. However, EG movement is not passive. Followers must interpret external cues, predict the partner’s movement, select an appropriate motor response, and execute that response in real time. Prior literature supports this interpretation: M1, PoCG/S1, SMA, striatum, cerebellum, IPL, and frontal regions are not exclusive to either IG or EG movement, but recur across self-paced, externally paced, rhythmic, and force-production tasks [50, 53, 56–61]. Thus, EG training may scaffold sensorimotor and cortico-striatal engagement by constraining the action plan through external information, rather than by eliminating striatal involvement. In other words, external cues may reduce the need to generate the full movement plan internally, while still requiring striatal-motor circuits to participate in cue-action mapping, response selection, sequencing, and execution.

The behavioral results support this more nuanced interpretation. Leaders and Followers both showed significant within-group improvements in UPDRS total and UPDRS Part III, and the pooled Dance group showed robust improvement on these global outcomes as well as significant improvement in the leg-related UPDRS subscore. Thus, the dance groups’ clinical improvement cannot be reduced to a single neural pattern. Leaders improved in the context of relative connectivity stability and possible normalization of disease-elevated cerebellar-sensorimotor connectivity, whereas Followers improved in the context of greater sensorimotor and caudate-motor connectivity change.

The disease-signature mapping further clarifies this distinction. Four significant trial pairs overlapped with the independently derived PwPD-vs-HOA disease-connectivity signature: PreCG/M1-CB Lobule VIIIb, left dorsomedial PoCG/S1-CB Lobule VIIIb, left dorsomedial PoCG/S1-insula, and insula-CB Lobule VIIIb. The two cerebellar-sensorimotor pairs were elevated in PwPD relative to HOA and appeared in the trial as post-intervention Controls > Leaders effects. By contrast, the within-Follower and difference-of-differences changes, including the caudate-dorsomedial PreCG/M1 edge, did not independently distinguish PwPD from HOA in the companion dataset. This means that Leaders’ pattern is more directly anchored to independently validated disease-relevant circuitry, whereas Followers’ pattern may reflect a different, intervention-specific form of sensorimotor recruitment.

The secondary Dance-vs-Control analysis is important in the context of arts-based interventions because it asks not whether the Leader or Follower roles are different or which one is more effective, but which neural pathways are engaged by participation in adapted tango as a whole, regardless of role, compared with a socially interactive education control condition. When Leaders and Followers were pooled, the resulting pattern was not simply a weaker version of the Follower findings. Instead, different networks emerged, including post-intervention Dance-vs-Control differences involving caudate, insula, precuneus, cerebellum, SMA, and IFG, as well as within-Dance pre-to-post changes involving sensory, motor and SMA-related regions. This finding suggests that the pooled dance intervention engaged both IG- and EG-relevant circuitry, consistent with the fact that adapted tango combines self-initiation, rhythmic entrainment, partner cueing, balance control, and whole-body motor coordination. Behaviorally, the pooled Dance group also showed significant improvement in the leg-related UPDRS subscore. This is clinically plausible because the leg subscore includes items related to lower-limb movement, gait, freezing, toe tapping, leg agility, posture, and lower-extremity tremor, all of which are directly challenged by adapted tango’s stepping patterns, weight shifts, pauses, turns, and multi-directional movements.

As a whole, the results suggest that dance-based training may support clinical improvement through more than one neural route. Leaders may benefit through relative stabilization, remediation or normalization of disease-elevated cerebellar-sensorimotor connectivity. Followers may benefit through cue-supported recruitment of sensorimotor, premotor, parietal, and caudate-motor circuitry. Pooled dancers showed a partly distinct network profile, suggesting that adapted tango as an arts- and dance-based intervention may engage a broader mixture of IG- and EG-weighted pathways than either role-specific contrast captures alone. Controls showed a partly overlapping pattern, with caudate- and sensorimotor-related increases, but without clinical improvement, emphasizing that increased connectivity is not necessarily beneficial in all cases. Its interpretation depends on the circuit, the group, and the behavioral context.

### Circuit-specific considerations

The cerebellum is central to the interpretation of the present findings, but its role should be understood in a region-specific rather than global way. Prior IG/EG neuroimaging studies consistently implicate the cerebellum in timing, coordination, error correction, and cue-based movement. Cerebellar activity has been reported during self-paced ankle or hand movements, self-generated finger key presses, and externally cued ankle movements, rhythmic tapping, finger tapping, hand force production, and phasic hand and foot movements [8, 12, 49, 50, 57, 60, 62–67]. This literature supports the broad role of cerebellar circuits in both internally and externally structured movement, while PD-specific studies suggest that cerebellar recruitment may become particularly important when basal-ganglia-mediated internal cueing is inefficient [8, 16, 49]. Our results tap into this literature in two ways: first, Controls showed increased post-intervention connectivity relative to Leaders in PreCG/M1-CB Lobule VIIIb and PoCG/S1-CB Lobule VIIIb, pairs that were also elevated in PwPD relative to HOA in the companion disease-signature analysis; second, Followers, who trained in the more EG-weighted role, showed greater pre-to-post change than Leaders in fusiform gyrus-CB Lobule VIIIa. Together, these findings suggest that cerebellar connectivity may index both disease-elevated compensatory circuitry in Controls and more task-specific cue-supported reorganization in Followers, depending on the connected target and analytic contrast.

The caudate is a second region warranting specific discussion. Classically, the caudate and related striatal structures are central to STC circuitry and to internally guided action selection. However, the literature reviewed in Theofanopoulou et al. (2026a) [17] indicates that striatal regions are not engaged only by IG tasks. Caudate/putamen activation has been reported during externally guided hand force production and finger tapping, rhythmic tapping, movement to metered beats, and externally cued ankle movements, as well as during self-paced or self-initiated tasks [12, 49, 51, 54–56, 68–70]. The understanding of this phenomenon is important for interpreting the present caudate-dorsomedial PreCG/M1 result. That Followers showed greater caudate-M1 change than Leaders does not contradict the EG framework. Instead, it suggests that EG training can still engage striatal-motor circuitry when external cues must be translated into selected, sequenced, and executed actions. The Follower role may reduce the need to internally generate the complete movement plan from scratch, but it does not remove the need for action selection. Rather, the Follower role shifts part of the action-selection process toward cue-action mapping, where tactile, proprioceptive, and auditory information constrain the motor response that the participant then generates.

The Control caudate result requires a different emphasis. Controls also showed greater caudate-dorsomedial PreCG/M1 change than Leaders, but without clinical improvement. Therefore, increased caudate-M1 connectivity cannot be interpreted as straightforwardly adaptive. It may reflect compensation, inefficient coupling, or broader motor-network hyperconnectivity. This interpretation also fits with the companion disease-signature results, in which caudate connectivity was clinically meaningful but target-specific: reduced lateral caudate-superior medial frontal connectivity was associated with greater disease severity as measured by the UPDRS-III, whereas the present caudate-M1 trial network was not itself one of the independently validated PwPD-vs-HOA disease-signature edges. Thus, the caudate appears to function as a clinically relevant hub, but the meaning of increased caudate connectivity depends on the connected target and the behavioral context.

M1 and S1 are also central to the present findings. Prior IG/EG studies repeatedly implicate PreCG/M1 and PoCG/S1 across self-paced finger tapping, ankle dorsiflexion, hand and foot movement, rhythmic tapping, and externally cued movement tasks [48, 52, 53, 56, 57, 71, 72]. In PD specifically, M1 and S1 effects can vary by task and disease context, with studies reporting both increased and decreased recruitment depending on whether the task is internally generated, externally cued, rhythmic, or performed under medication-related conditions [16, 51, 71]. This variability helps explain why M1-related effects in the present trial should not be interpreted simply as “more is better” or “less is better.” Leaders’ lower M1-cerebellar connectivity relative to Controls may reflect reduced disease-like hyperconnectivity on a validated edge, whereas Followers’ increased caudate-M1 and S1-M1 connectivity may reflect cue-supported sensorimotor integration.

Further, the effector-specific organization of M1 provides useful context for interpreting the behavioral findings. The present trial’s M1 components were not broad undifferentiated motor regions: as shown in Theofanopoulou et al. (2026a) [17], dorsomedial PreCG/M1 components (IC54) overlap leg- and hand-related motor zones, while dorsolateral PreCG/M1 (IC66) overlaps hand- and larynx-related zones. For the former (M1 leg/hand area), Followers and Controls showed a greater pre-to-post increase than Leaders in with the lateral caudate. For the former (M1 hand/larynx area), Followers showed increased connectivity with right dorsolateral PoCG/S1 in within-group comparisons, while the pooled Dance group showed pre-to-post decreases in connectivity with right dorsolateral PoCG/S1, left dorsomedial PoCG/S1, and SMA. These M1 patterns are potentially relevant to the behavioral improvements because the UPDRS measures that improved in Leaders and Followers encompass multiple motor effectors, including upper- and lower-limb function as well as laryngeal-related motor function (e.g., speech, swallowing). The involvement of dorsolateral PreCG/M1 (hand/larynx area) is particularly interesting in light of recent work in PD using more sensitive automated speech-performance measurements showing that a dance-based intervention can improve vocal and prosodic measures [73]. Future studies incorporating specialized vocal tasks and quantitative speech measures may therefore be better positioned to test whether dance-based IG/EG training modulates speech performance and laryngeal motor circuitry.

The insula and precuneus also merit careful interpretation. Although they are not as central to the classic IG/STC versus EG/CTC distinction as M1, caudate, or cerebellum, both regions appear in prior IG/EG movement studies. Insula recruitment has been reported during self-paced finger tapping and self-initiated grasping [74, 75], while precuneus involvement has been reported during self-initiated grasping, self-initiated tracing, self-paced ankle movements, and paced finger tapping [48, 58, 61, 75, 76]. IPL, which appeared in the Follower > Leader difference-of-differences pattern, has likewise been implicated in ankle, foot and finger movements, and self-paced phasic hand and foot movements [12, 53, 61]. These regions may therefore reflect broader sensorimotor integration, attention to body position, action monitoring, or transformation of sensory cues into movement plans rather than belonging exclusively to either the IG or EG pathway.

Taken together, the circuit-specific findings support a model in which the intervention did not simply increase STC connectivity in Leaders and CTC connectivity in Followers. Instead, Leaders showed clinical improvement with relative rsFNC stability and possible remediation of disease-elevated cerebellar-sensorimotor circuitry. Followers showed clinical improvement with distributed sensorimotor, premotor, parietal, caudate-motor, and limited cerebellar recruitment. Controls showed overlapping caudate- and sensorimotor-related changes without behavioral benefit. This pattern suggests that IG- and EG-weighted dance roles may engage different forms of motor-network adaptation, with normalization and compensation operating as partially complementary rather than mutually exclusive mechanisms.

### Limitations

This study has several limitations. First, our interpretive framework is grounded in prior literature identifying specific brain regions as being associated with IG movement, EG movement, or both. Accordingly, we focused on a predefined set of ROIs rather than conducting a whole-brain analysis. This approach allowed direct comparison with the companion PwPD-vs-HOA study and enabled precise mapping of the present trial’s significant edges onto the same NeuroMark IC framework. However, it also means that our findings are limited to the connectivity patterns observed within the selected ROIs and their corresponding NeuroMark ICs. Other regions or finer-scale dynamics relevant to IG/EG movement control may not have been captured. Second, the leader/follower manipulation should be interpreted as a relative IG/EG weighting rather than as a categorical separation of neural systems. Leaders self-initiate direction, timing, and amplitude, but they also respond to music, spatial context, and partner feedback. Followers rely more heavily on auditory, tactile, and proprioceptive cues, but they must still generate internally coordinated motor responses. Thus, Leaders are not purely IG and Followers are not purely EG. This overlap may attenuate between-role differences and may partly explain why both dance groups improved behaviorally while showing different, but not strictly opposite, rsFNC profiles. Third, none of the rsFNC findings survived correction for multiple comparisons across the full set of tested IC pairs. The rsFNC results were therefore interpreted at the uncorrected p<0.01 threshold. Although these patterns are anatomically coherent and consistent with the a priori IG/EG framework, they require replication in larger samples. Fourth, the disease-signature trajectory analysis should be treated as exploratory. The mapping to the companion PwPD-vs-HOA dataset is a strength because it allowed significant trial edges to be interpreted relative to independently identified disease-related connectivity patterns. However, the HOA reference sample was cross-sectional and was not part of the randomized trial. The trajectory analysis therefore provides directional context, not definitive evidence of normalization. Despite these limitations, the present findings provide an anatomically grounded account of how adapted tango may influence motor networks in PD.

## Data availability

The authors confirm that, for approved reasons, some access restrictions apply to the data underlying the findings. Public data deposition is not ethical or legal and would compromise patient privacy. Data are, in part, housed in the VINCI Veterans Affairs (VA) Informatics and Computing Infrastructure (VINCI), a secure, virtual computing environment developed through a partnership between the VA Office of Information Technology (OI&T) and the Veterans Health Administration’s Office of Research and Development (VHA ORD). Data are also stored on a VA secured server under a Data Usage Agreement with Emory University. VINCI is a partner with the Corporate Data Warehouse (CDW) and hosts all data available through CDW. VHA National Data Services (NDS) authorizes research access to patient data. Data are only available for researchers who meet the criteria for access to confidential data. Contact: ^35^. Detailed analysis and results files of aggregated data can be found at https://github.com/nehabajaj101/Adaptive-Tango-Intervention-/tree/main.

## Acknowledgements

We thank the volunteers and participants for their time and effort devoted to this study.

